# Family planning uptake and its associated factors among women of reproductive age in Uganda: an insight from the Uganda Demographic and Health Survey 2016

**DOI:** 10.1101/2022.08.31.22279440

**Authors:** Anthony Mark Ochen, Primus Che Chi

## Abstract

Despite the government efforts to reduce the high fertility levels and increase uptake of family planning services in Uganda, family planning use was still low at 30% which is the lowest in the East African region. This study was undertaken to determine the prevalence and factors associated with the uptake of family planning methods among women of reproductive age in Uganda. This is a community-based cross-sectional study that utilized secondary data from the Uganda Demographic and Health Survey (UDHS) of 2016. The survey data was downloaded from the Measure Demographic Health Survey website after data use permission was granted. Data was collected from a representative sample of women of the reproductive age group (15-49 years) from all the 15 regions in Uganda. A total of 19,088 eligible women were interviewed but interviews were completed with 18,506 women. Data analysis was performed using SPSS statistical software version 32.0 where univariable, bivariable, and multivariable analyses were conducted. The prevalence of family planning use was found to be 29.3% and that of modern contraceptive use was found to be 26.6%. Multivariable analysis showed higher odds of current family planning use among older women (40-44 years) (aOR=2.09, 95% CI: 1.40-3.12); women who had attained the secondary level of education (aOR=1.91, 95% CI: 1.32-2.76); those living in households with the highest wealth index (aOR=1.87, 95% CI: 1.29-2.72); and awareness of the availability of family planning methods (aOR=1.41, 95% CI: 1.17-1.72). In conclusion, the study found a low prevalence of current family planning use among women of reproductive age and this calls for more efforts toward the advancement of women’s education, awareness creation, designing policies to popularize use of FP among adolescents and young women, and socio-economic empowerment of women.

## Introduction

Uganda has one of the fastest-growing populations in the sub-Saharan Africa (SSA) region at a rate of 3.2% per annum (1). It has a persistently high fertility rate of 5.4 children born per woman which is higher than the total wanted fertility rate of 4.3 (2). The use of family planning (FP) among currently married women increased from 23% in 2000 to 39% in 2016, however, the increase was most pronounced for the use of modern methods which rose from 18% in 2001 to 35% in 2016 (2). According to the Uganda Demographic and Health Survey (UDHS) 2016 estimates, there were 336 maternal deaths per 100,000 live births and an infant mortality rate of 43 deaths per 1,000 live births (2). This poses a great threat to the development and well-being of the Ugandan population as reflected in the high infant mortality rates and maternal mortality ratios. High birth rates not only affect maternal and child mortality but frustrate governments’ efforts in the provision of social and health services to communities.

The World Health Organisation (WHO) refers to family planning (FP) as a process that allows people to attain their desired number of children and determine the spacing of pregnancies, which is achieved through the use of FP methods and treatment of infertility (3). Of the 1.9 billion women of reproductive age group (15-49 years) worldwide in 2019, 1.1 billion needed FP methods; of these, 842 million were using contraceptive methods, and 270 million had an unmet need for contraceptives (4). The modern contraceptive prevalence among married women of reproductive age increased worldwide between 2000 and 2019 by 2.1% from 55.0% to 57.1% (4). The explanations for this slow increase include the limited choice of methods; limited access to services particularly among young, poorer, and unmarried people; fear or experience of side effects; cultural or religious opposition; poor quality of available services; users and providers bias against some methods; and gender-based barriers to accessing services (4).

The United Nations (UN) estimates the total fertility rate (TFR) of the sub-Saharan Africa region at 4.7 births per woman in 2015-2020 which is more than twice the level of any other world region (5). Consequentially, the population of sub-Saharan Africa is expected to grow from 1 billion in 2015 to about 2 billion in 2050 and nearly 4 billion in 2100 (5). Therefore, universal access to FP services is an important global strategy to control fast-growing populations and improve maternal and child health. There are great benefits of investing in FP including reduced maternal and neonatal mortality through decline in abortions and pregnancies (6). However, several developing countries including Uganda stepped into the post-millennium development goals (MDG) era with a national health profile that required substantial improvements, including access to FP services (7, 8). Furthermore, in 2019, over 190 million (10%) of married women were estimated to have an unmet need for FP where the prevalence was higher in Africa compared to other parts of the world (9). This is well articulated in the sustainable development goals (SDG) 3, target 3.7 calls on countries “by 2030, to ensure universal access to sexual and reproductive health-care services, including for FP, information and education, and the integration of reproductive health into national strategies and program”; with specifically 3.7.1 which calls for universal access to FP services to ensure healthy lives and well-being (10).

Despite the government efforts to reduce high fertility levels and increase uptake of FP services in Uganda, the prevalence rate is only 30% among married women which is the lowest in the East African region. Factors contributing to the low FP rates are multi-factorial and includes; limited accessibility to contraceptives, long distance to the health facility, few qualified health experts, fear of side effects, limited male involvement, religion or cultural beliefs, polygamous marriage, and lack of awareness (11-16). Monitoring factors influencing the uptake of FP services is important to target scarce public resources to those with more need and enhance the progress towards achieving the global targets. Thus far, a recent study that has been published in Uganda only focussed on factors associated with modern contraceptives among female adolescents (17). Therefore, this study was undertaken to examine the prevalence and factors associated with the current FP uptake among women of reproductive age in Uganda using the 2016 Uganda Demographic and Health Survey data.

## Methods and Materials

### Ethics statement

The survey was approved by the Uganda National Council for Science and Technology (UNCST). Respondents were informed about the survey and written informed consent was obtained from the parents/guardian of each participant under 18 years of age. The authors received the survey data from the USAID DHS program database after a request to download the dataset was granted. After data access was authorized, the authors of this study maintained the confidentiality of the dataset.

### Study context

This study utilized secondary data from the Uganda Demographic and Health Survey (UDHS) 2016. The UDHS 2016 is a part of the global program implemented by the Uganda Bureau of Statistics (UBOS) in collaboration with the Ministry of Health (MoH). The funding for the UDHS 2016 was provided by the Government of Uganda, the United States Agency for International Development (USAID), the United Nations Children’s Fund (UNICEF), and the United Nations Population Fund (UNFPA). The DHS is undertaken every five years and the 2016 survey is the sixth DHS in Uganda, the first one was conducted in 1988.

To generate statistics that were representative of the country as a whole in the 15 regions, the number of women surveyed in each region contributed to the size of the total sample in proportion to the size of the region. This is because some regions had small populations and others had large populations. The 15 regions of Uganda where the UDHS 2016 was implemented were; South-Central, North-Central, Kampala, Busoga, Bukedi, Bugisu, Teso, Karamoja, Lango, Acholi, West-Nile, Bunyoro, Tooro, Kigezi, and Ankole Regions (Fig 1).

**Figure 1.**
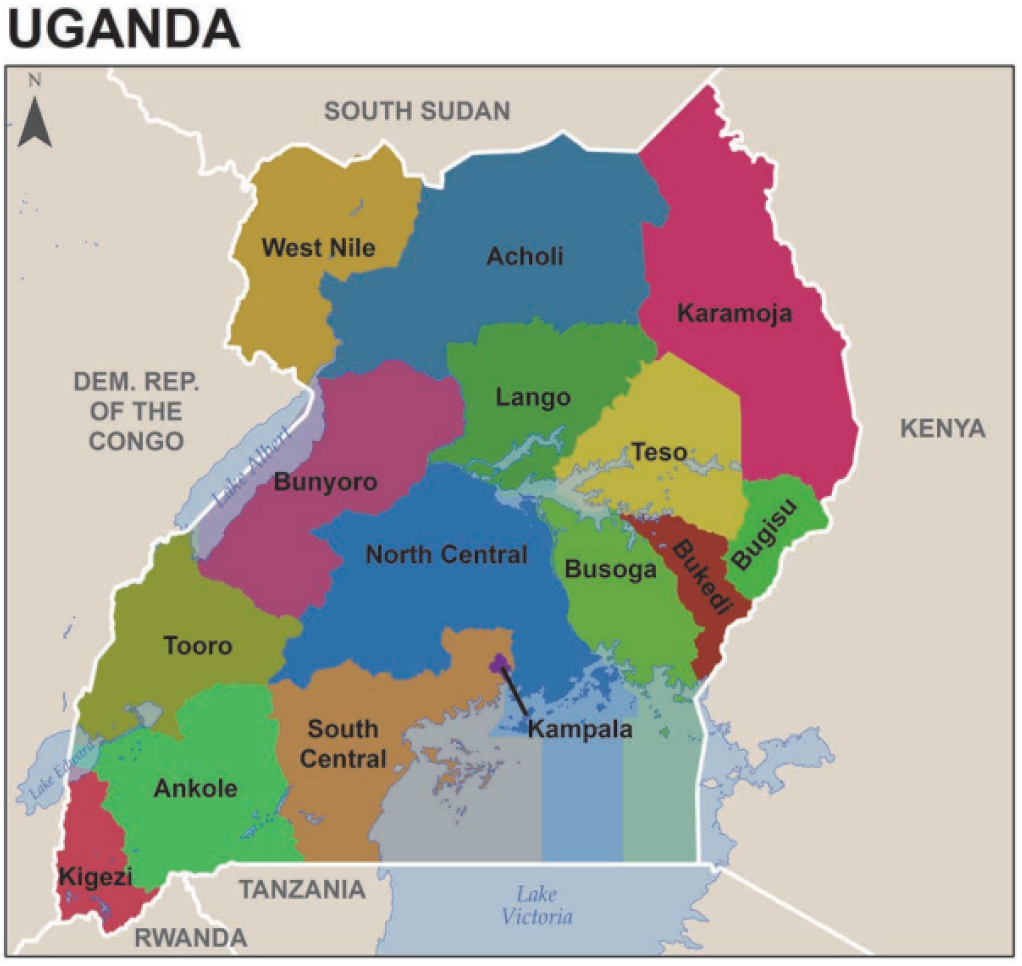
Map of Uganda showing 15 regions where the 2016 UDHS was implemented. From the 2016 UDHS Main Report

### Study design

This was a community-based cross-sectional study where data was collected from a representative sample of women of the reproductive age group (15-49 years).

### Target population

The study participants were all women in the reproductive age group of 15-49 years living in Uganda at the time of the survey.

### Sample size

A national representative sample of 20,880 households was selected for the study. From these households, a total of 19,088 eligible women in the reproductive age group were interviewed using a structured questionnaire (18). However, interviews were completed with 18,506 women, yielding an overall response rate of 97%. Response rates were higher in rural (97.6%) than in urban areas (94.8%).

### Sampling procedure

The UDHS 2016 used a multi-stage stratified sampling method (in two stages) to select the study participants. Three regions (South Central, North Central, and Busoga) were stratified into island and non-island sub-regions. Each region/sub-region was stratified into urban and rural areas yielding 34 sampling strata. In the first stage, 697 Enumeration Areas (EA) were selected, 162 EA in urban and 535 in rural areas (Fig 2). One cluster was eliminated due to disputed boundaries leaving a total of 696 clusters. Households constituted the second stage of sampling. A listing of households was compiled in each of the 696 EAs from April to October 2016. The listing excluded institutional living arrangements such as army barracks, hospitals, police camps, and boarding schools. Only one segment was selected for the survey with probability proportional to size, and the household listing was conducted only in the selected segment. In total, a representative sample of 20,880 households was randomly selected for the UDHS 2016. All women aged 15-49 who were either permanent residents of the selected households or visitors who stayed in the household the night before the survey were interviewed.

**Figure 2.**
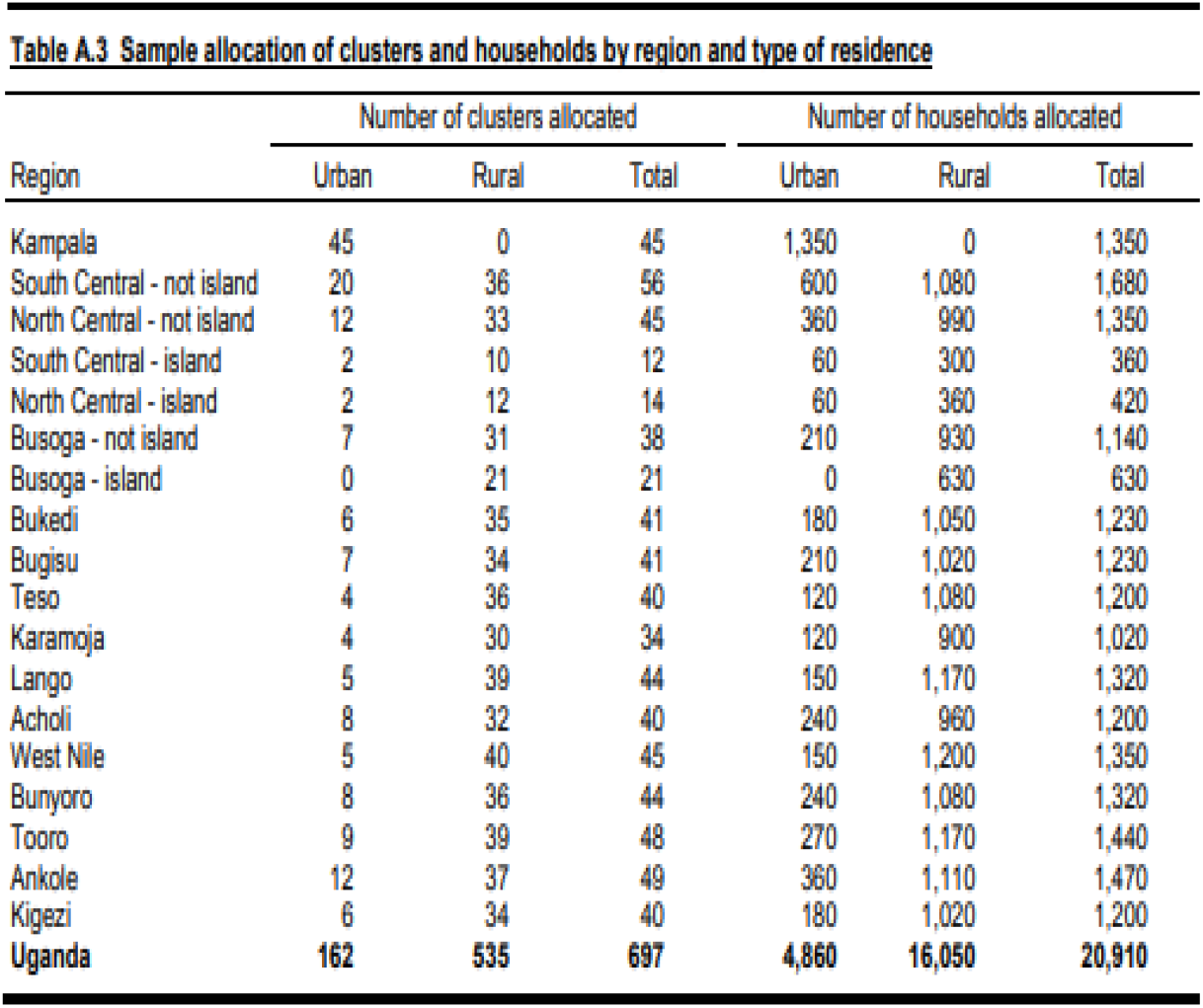
From the 2016 UDHS Main Report.

Due to the non-proportional allocation of the sample to different regions and their urban and rural areas, and the possible differences in response rates, sampling weights were analysed using the UDHS 2016 data to ensure that the survey results were representative at the national level as well as at the regional and sub-regional levels. After adjusting for non-response, the sampling weights were normalized to get the final standard weights that appeared in the data files. The normalization process was aimed at obtaining a total number of un-weighted cases equal to the total number of weighted cases using normalized weights at the national level, for the total number of households and women.

### Data collection procedure

A structured and pre-tested questionnaire was used as a tool for data collection. The questionnaire was developed in English and then translated into nine different local languages. The questionnaire was developed based on standard DHS survey questionnaires and programmed into tablet computers to facilitate computer-assisted personal interviewing (CAPI) for data collection purposes, with the capability to choose any of the nine languages for each questionnaire. The UDHS and ICF technical teams trained 45 participants who administered the paper and electronic questionnaires with tablet computers. All trainees had some experience with household surveys. The technical teams conducted debriefing sessions with the pre-test field staff and modifications to the questionnaires were made based on lessons learned from the exercise. A total of 173 fieldworkers (108 women and 65 men) were recruited and trained to serve as supervisors, CAPI managers, interviewers, health technicians, and reserve interviewers for the main fieldwork. The training course included instruction on interviewing techniques and field procedures, a detailed review of questionnaire content, instruction on administering the paper and electronic questionnaires, mock interviews between participants in the classroom, and practice interviews with actual respondents in areas outside the 2016 UDHS sample. A two-day field practice was organized to provide trainees with additional hands-on practice before the actual fieldwork.

A total of 84 participants were selected to serve as interviewers, 21 as health technicians, 21 as field data managers, and 21 as team leaders. The selection of team leaders and field data managers was based on experience in leading survey teams and performance during the pre-test and main training. Supervisory activities included assigning households and receiving completed interviews from interviewers, recognizing and dealing with error messages, receiving system updates and distributing updates to interviewers, resolving duplicated cases, closing clusters, and transferring interviews to the central office via a secure Internet file streaming system (IFSS). Data collection was conducted by 21 field teams, each consisting of one team leader, one field data manager, three female interviewers, one male interviewer, one health technician, and one driver. Electronic data files were transferred from each interviewer’s tablet computer to the team supervisor’s tablet computer every day. The field supervisors transferred data to the central data processing office via IFSS. Senior staff from the Makerere University School of Public Health, the Ministry of Health, and UBOS, and a survey technical specialist. The DHS Program coordinated and supervised fieldwork activities. Data collection took place from 20 June 2016 through 16 December 2016.

### Variables and measurements

The outcome variable of our study is the use of current family planning methods (traditional, folkloric, and modern methods) among women of reproductive age. For this study, we used the women’s questionnaire which collected information from women of reproductive age, 15-49 years. The women’s questionnaire consisted of 12 sections, however, we used variables for four sections; section 1 – respondent’s background, section 3 – contraception, section 9 – fertility preferences, and section 10 – husband’s background and woman’s work. The independent variables used were: age, place of residence, level of education, literacy level, current marital status, religious affiliation, current employment, husband’s education, wealth index, ethnicity, regions in Uganda, knowledge of family planning methods, source of information (radio, television, newspaper, or text message via phone), decision-maker for using contraceptives, last source for FP users, visited health facility and being visited by a field worker in the last 12 months, and whether participants were told about other FP methods at the health facility.

### Data management and analysis

The downloaded data was entered into the SPSS software version 32.0 and data was cleaned, transformed to populate cells with few values, and re-coded as well. The collinearity effect was checked using a cut-off of the value of variance inflation factor (VIF) less than 6. The univariable, bivariable, and multivariable analyses were performed. The Univariable analysis was used to summarise the socio-demographic factors to find the pattern within the dataset meanwhile, the bivariable analysis was used to compare two variables to measure the relationship between them, and also identify variables to include in the regression analysis. On the other hand, multivariable analysis, which is a more complex analysis technique was used to understand interactions between two or more variables and also control for confounding factors. At the univariable level, frequencies and proportions were determined. At the bivariable level, analysis was done by cross-tabulation using Pearson Chi-Square (x^2^) test for categorical variables. Pearson’s Chi-square test was used because it is appropriate to analyse data with a binary outcome and independent categorical variables.

The associations between the outcome and independent variables were measured using the odds ratio (OR) for which a 95% confidence interval (CI) was calculated. All variables that showed a significant association of p<0.05 at the bivariable level were further analysed at the multivariable level using a binary logistic regression. Binary logistic regression analysis was used because the data set is normally distributed and has a binary outcome. Some variables were also included in the multivariable model because these variables showed influence on the outcome variable, hence we needed to identify whether each of them had been confounded by another variable or not. The adjusted odds ratio (aOR) was undertaken using a simultaneous modelling technique to determine the presence of associations between the outcome and independent variables. Model fitness was performed using Hosmer and Lemeshow Chi-Square test at p>0.05.

## Results of the Study

### Description of socio-demographic characteristics

A total of 18,506 samples of women of reproductive age (15-49 years) were included in the dataset where 23.1% were adolescents (15-19 years) and 76.3% lived in rural areas (Table 1). More than half (58.9%) attained primary level of education, 31.4% were married, and 40.8% were affiliated with Catholic religion. Further analysis revealed that majority of women (73.9%) were currently working at the time of the interview and most of them (21.8%) lived in households with highest wealth index. The Baganda tribe represented the highest proportion of ethnic groupings (13.2%) and only a handful of women (15.5%) used government clinic/pharmacy as the main source for family planning methods.

**Table 1.**
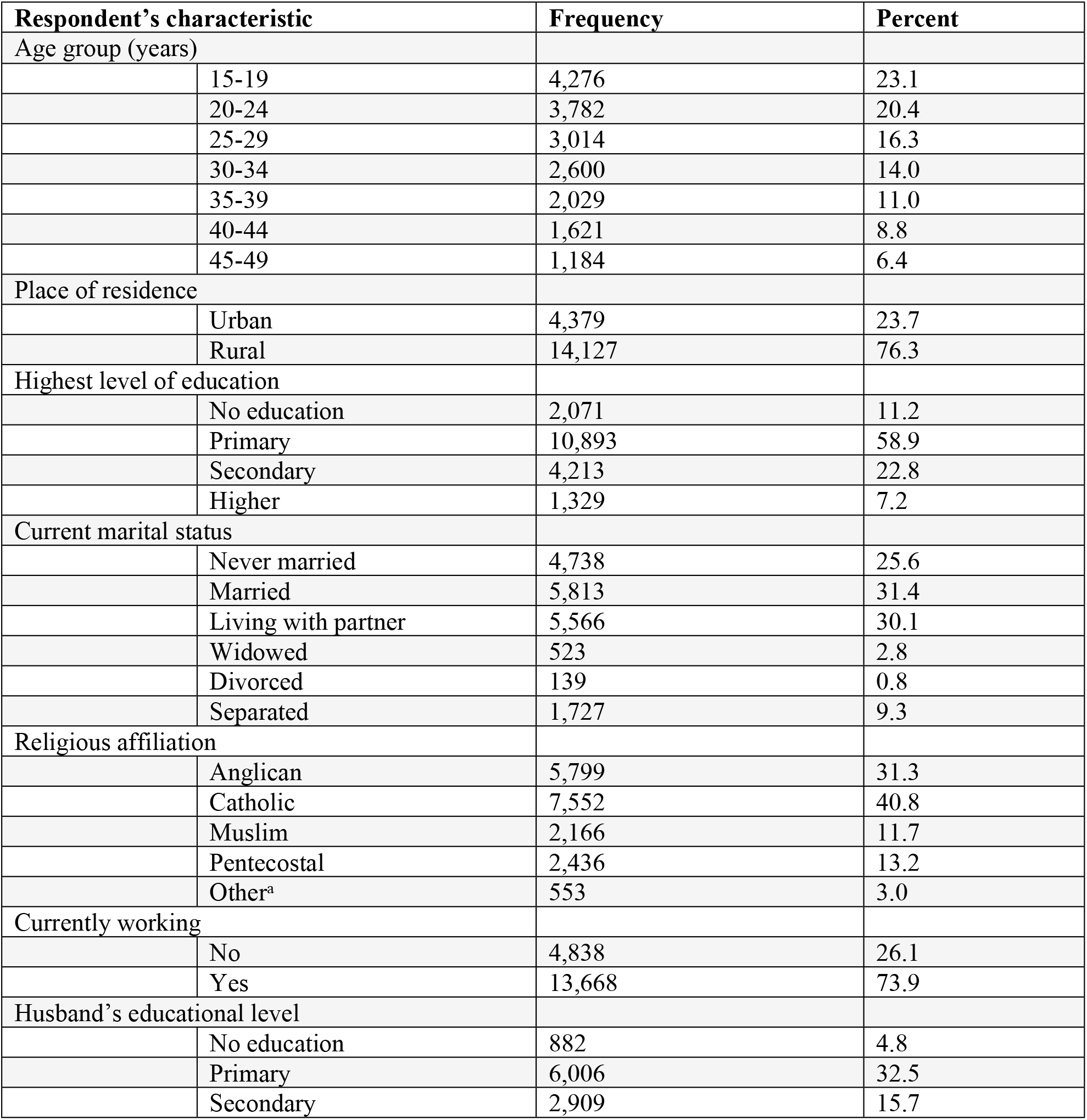
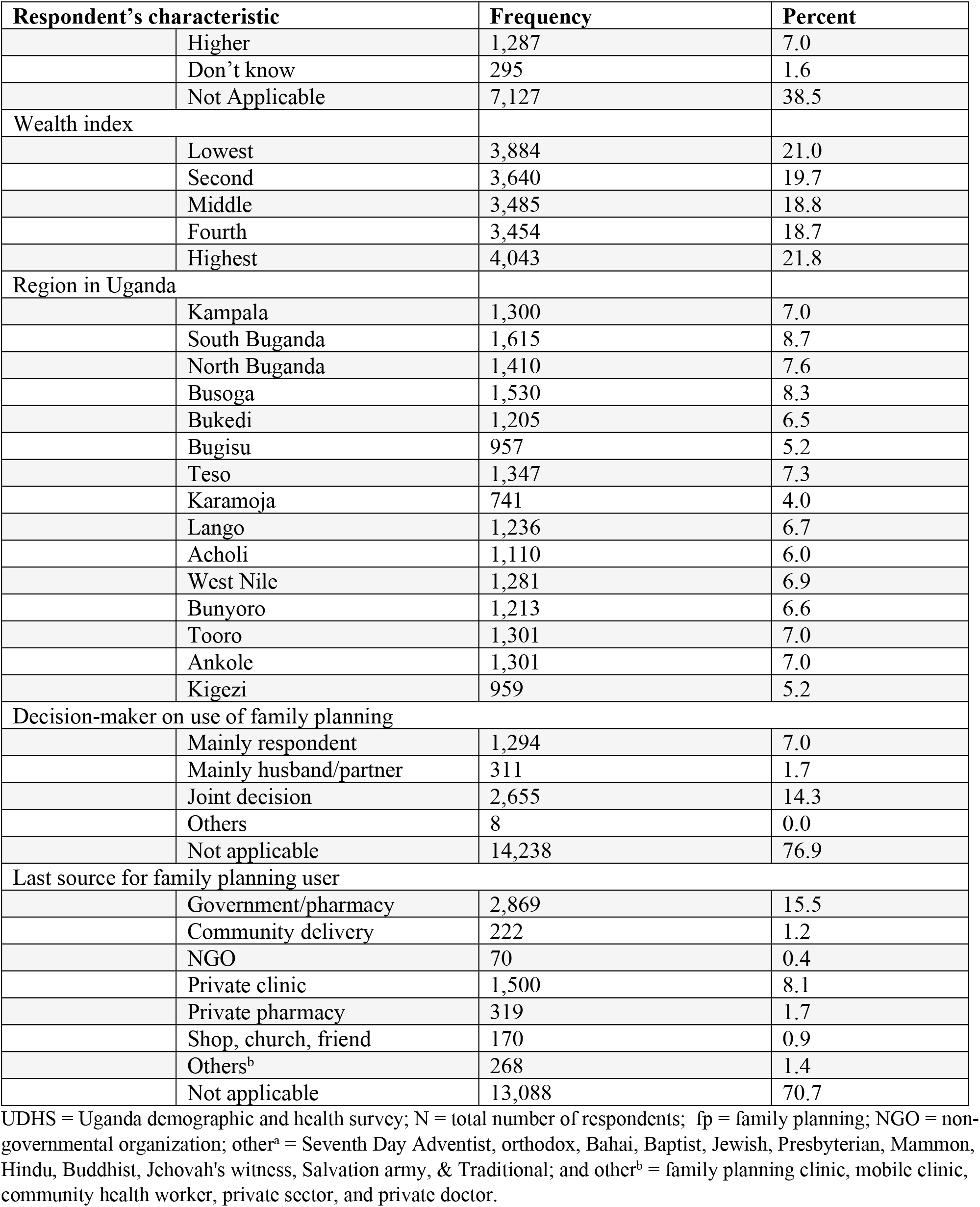
Socio-demographic characteristics of women of reproductive age, UDHS 2016 (N = 18,506)

### Prevalence of family planning uptake

The prevalence of current FP use among women was 29.3%, with 26.6% of them using modern contraceptive methods (Table 2). Most women preferred injections (13.5%), followed by implants (4.8%) and male condoms (2.9%). Analysis of the pattern of FP use showed that less than half (29.3%) were currently using at least one method, 12.1% used since last birth, 14.1% used before last birth, and 44.3% never used any FP method.

**Table 2.** Prevalence of current family planning uptake among women of reproductive age, UDHS 2016.

| Respondent characteristics |  | Frequency | Percent |
| --- | --- | --- | --- |
| Current use of FP methods |  |  |  |
|  | No | 13,088 | 70.7 |
|  | Yes | 5,418 | 29.3 |
| Current use by methods |  |  |  |
|  | No method | 13,088 | 70.7 |
|  | Folkloric method | 46 | 0.2 |
|  | Traditional method | 458 | 2.5 |
|  | Modern method | 4,914 | 26.6 |
| Current method type |  |  |  |
|  | Not using any method | 13,088 | 70.7 |
|  | Pill | 256 | 1.4 |
|  | IUD | 197 | 1.1 |
|  | Male condom | 534 | 2.9 |
|  | Female sterilization | 355 | 1.9 |
|  | Periodic abstinence | 169 | 0.9 |
|  | Withdrawal | 289 | 1.6 |
|  | Implant/Norplant | 858 | 4.8 |
|  | Lactational amenorrhea | 108 | 0.6 |
|  | Injections | 2,506 | 13.5 |
|  | Other | 116 | 0.6 |
| Pattern of family planning use |  |  |  |
|  | Currently using | 5,418 | 29.3 |
|  | Used since last birth | 2,247 | 12.1 |
|  | Used before last birth | 2,635 | 14.1 |
|  | Never used | 8,206 | 44.3 |
| Intention to use contraceptive |  |  |  |
|  | Using modern method | 4,914 | 26.6 |
|  | Using traditional method | 504 | 2.7 |
|  | Non-user intends to use later | 7,879 | 42.6 |
|  | Does not intend to use | 5,209 | 28.1 |

### Determinants of family planning uptake

The table of unadjusted and adjusted analyses is shown below (Table 3). The unadjusted analysis shows significantly higher odds of current FP use among; women who knew about FP methods (OR=7.44, 95% CI: 5.89-9.40); women whose husbands had attained a higher level of education (OR=3.29, 95% CI: 2.71-4.00); women who had attained a secondary level of education (OR=2.49, 95% CI: 2.14-2.90); those who had lived in households with highest wealth index (OR=2.17, 95% CI: 1.96-2.41); those who were in the age group of 25-29 years (OR=2.18, 95% CI: 1.87-2.55); and married women (OR=1.59, 95% CI: 1.42-1.79) as detailed in table 3.

**Table 3.**
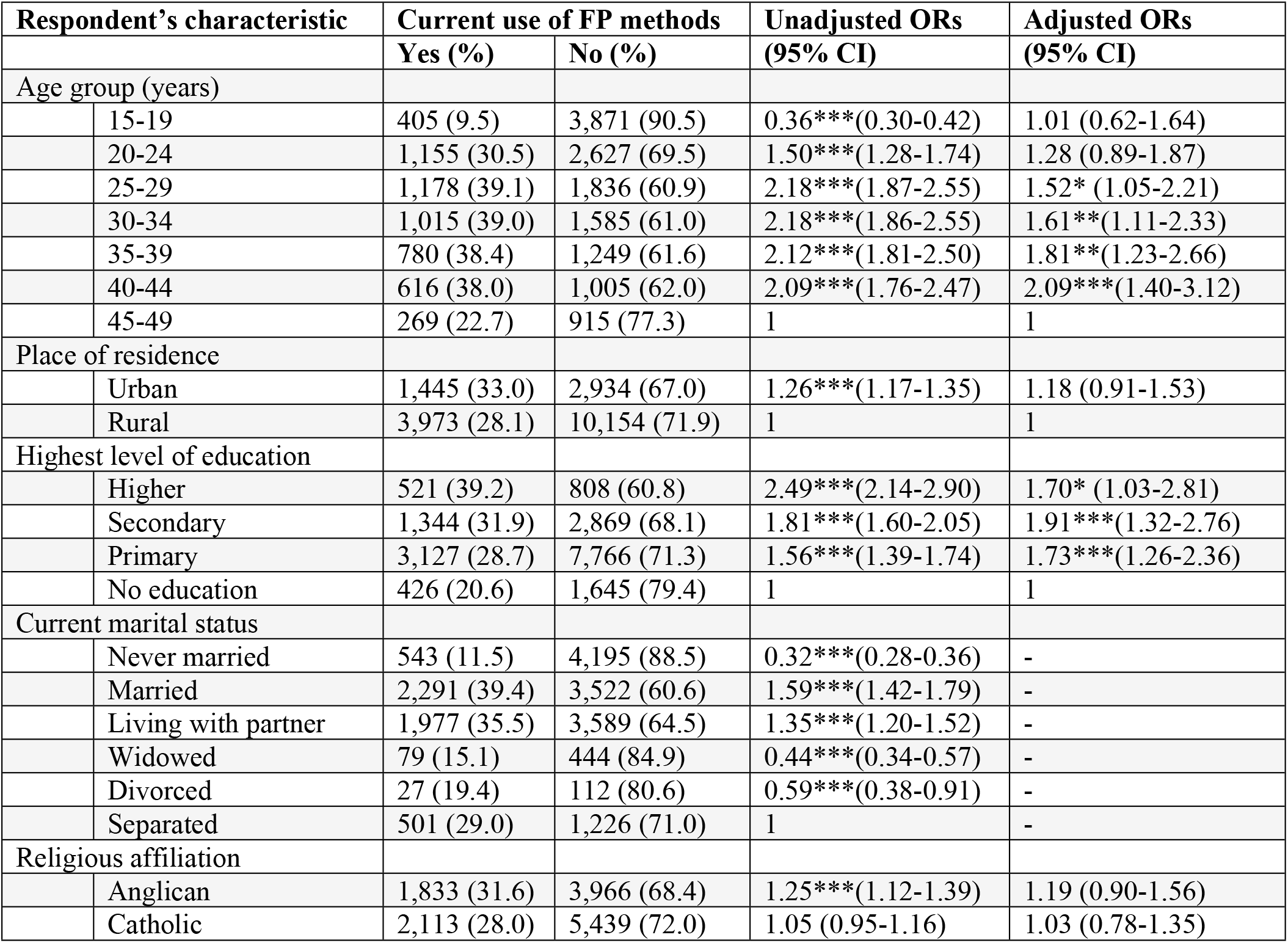
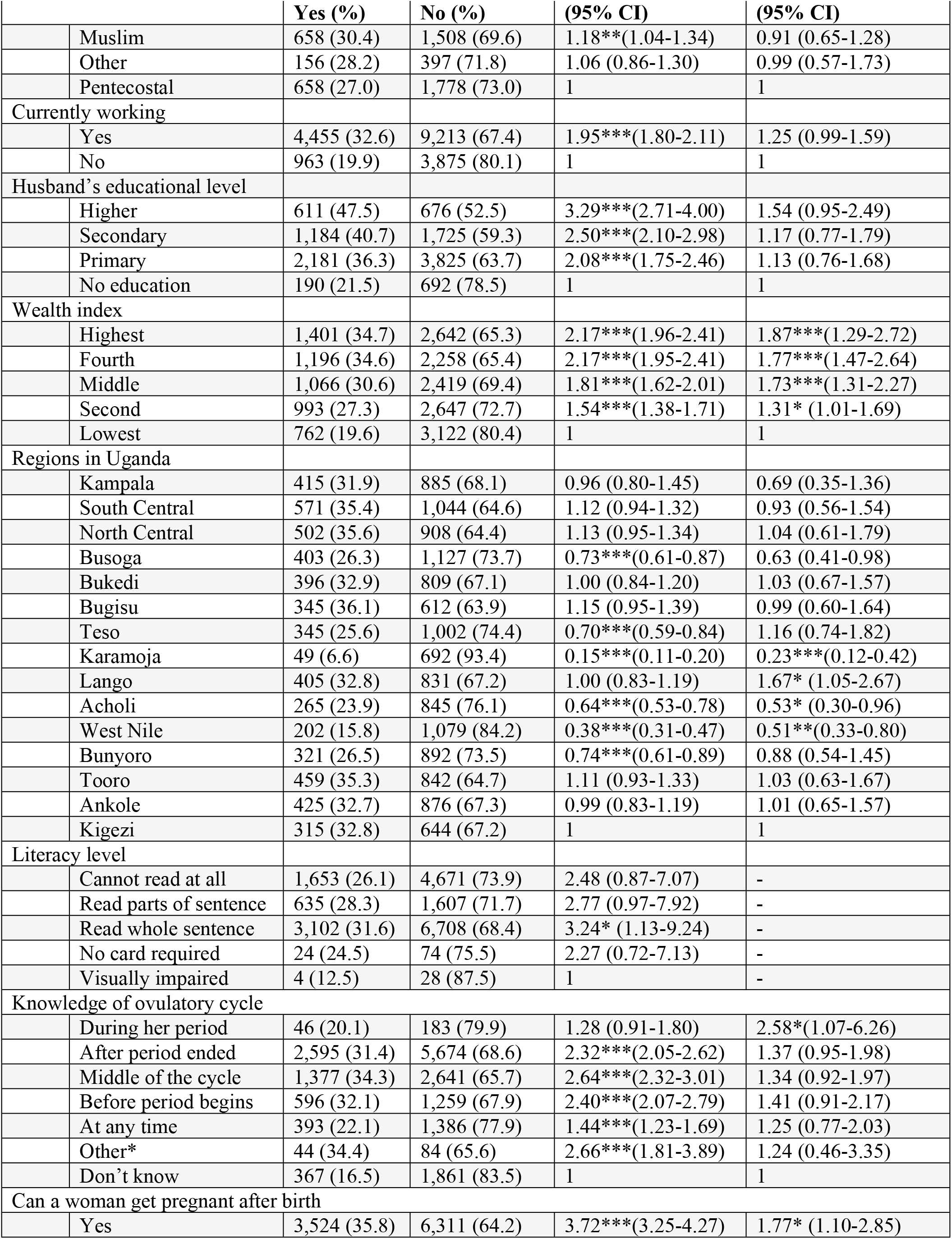
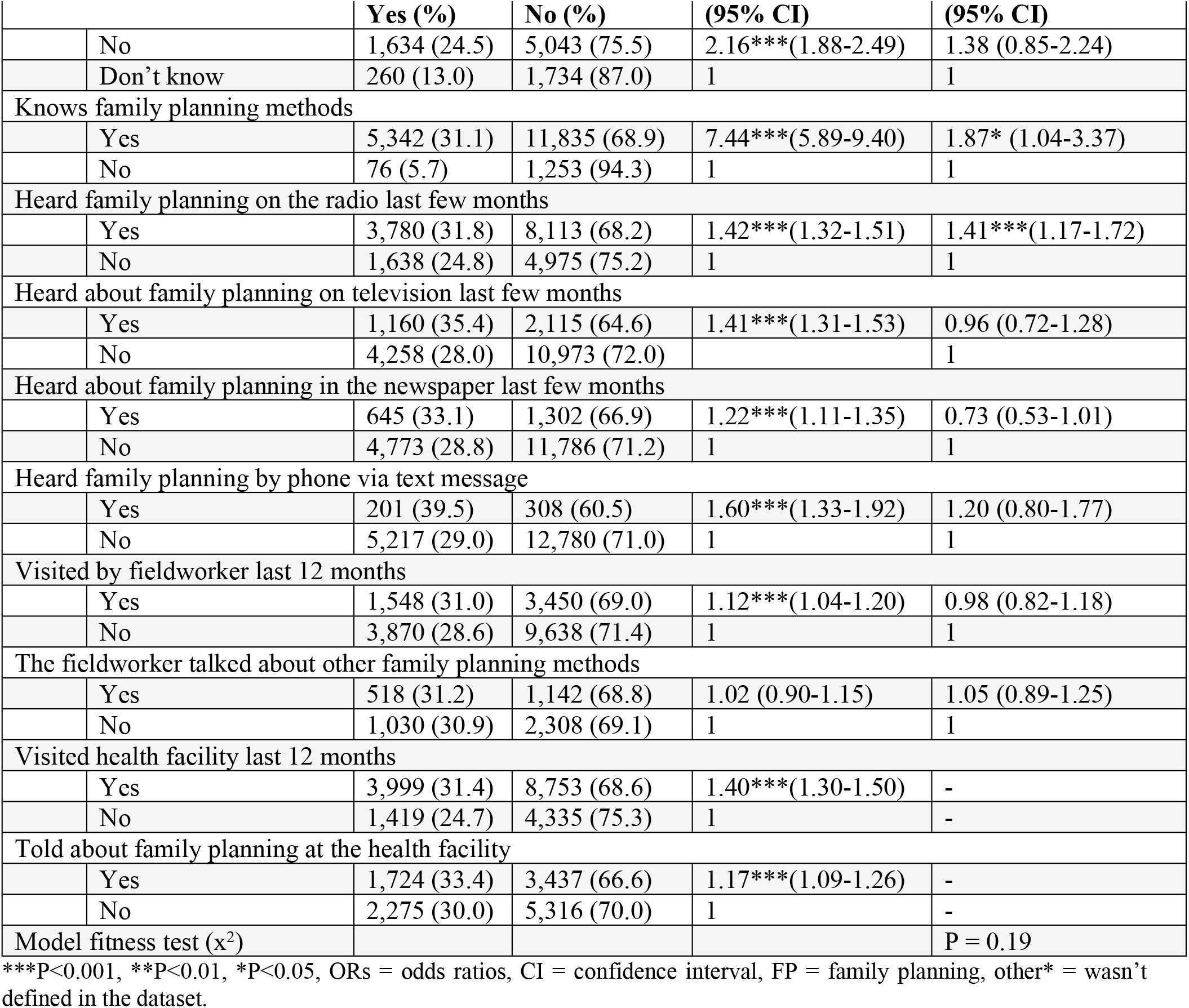
Unadjusted analysis of current family planning uptake among women of reproductive age.

The adjusted analysis revealed significantly higher odds of current FP use among; women who were knowledgeable about their ovulatory cycle (aOR=2.58, 95% CI: 1.07-6.26); Older women of age group 40-44 years (aOR=2.09, 95% CI: 1.40-3.12); women who had attained a secondary level of education (aOR=1.91, 95% CI: 1.32-2.76); those who lived in households with the highest wealth index (aOR=1.87, 95% CI: 1.29-2.72); those who were aware of the availability of FP methods (aOR=1.87, 95% CI: 1.04-3.37); and women living in the Lango sub-region of Uganda (aOR=1.67, 95% CI: 1.05-2.67). The overall model shows a good fit of data with the Pearson Chi-Square test of p = 0.19 and the model is said to fit well when the p-value is more than 0.05.

## Discussions

### Introduction

This study examined the prevalence and factors associated with the uptake of current FP methods among women of reproductive age (15-49 years) in Uganda. The study found the current family planning uptake prevalence to be 29.3%, which is an improvement from 24% in the UDHS 2011 (19). However, there was a decrease of 3.4% in the use of modern contraceptives from 30% in the UDHS 2011 to 26.6% in the UDHS 2016 (2, 19).

### Discussions of key results

Uganda has the lowest FP prevalence rate as compared to the rates in neighbouring countries like Kenya (45.5%), Rwanda (51.6%), and Tanzania (34.4%) (20). The difference in the FP prevalence rates could be due to the low level of education among women, having three or more children, living in rural areas, husband’s disagreement on contraceptive use, perceived side effects, infant mortality; negative traditional practices, knowledge gaps on contraceptive methods, fears, rumours, and misconceptions about specific methods and unavailability and poor quality of services (21). A recent study on contraceptive use further alluded to cultural beliefs, financial constraints to access contraceptives and limited sources of family planning information like television and newspapers (17). The low prevalence of family planning use could negatively affect Uganda’s progress in achieving sustainable development goal (SDG) 3 target 3.7 aimed at ensuring universal access to sexual and reproductive healthcare services, including family planning by 2030 if immediate interventions are not put in place (22).

Our study found a significantly higher odds of FP use among older women in the age group 40-44 years. The previous analysis of the 2011 UDHS and Uganda FP costed implementation plan 2015-2020 revealed disparities in the use of family planning by; age, marital status, education, socio-economic status, and rural-urban geographic location (23). Our finding concurs with other studies that found the use of FP increases with older age (24, 25). It is believed that older women are more exposed to information concerning childbearing and the dangers of high parities so they have appreciated the importance of the uptake of family planning methods.

Educational achievements of both women and their husbands were found in some studies to be very significant factors in the use of FP methods (25, 26). However, the present study found the education of women alone as the driver of the FP uptake. Unlike women with no formal education, women with at least secondary education are more likely users of FP methods (27). This does not come as much of a surprise as higher education attainment increases female decision-making powers and awareness of the benefits of good family planning practices. This affirms the relevance of education in matters concerning the use of FP in Uganda. Thus, the dispute in Uganda that universal secondary education (USE) works towards enhancing FP use is highly supported.

In our study, women living in households with the highest wealth index had significantly higher odds of FP. This finding concurs with a study conducted in Ethiopia (28) but, contradicts another study conducted in Uganda which found that wealth was not associated with FP use (29). This variation can be explained by current women’s empowerment through education and media awareness. The poor are less likely to be well informed about FP methods which can be attributed to a lack of ownership of television sets, mobile phones, or buying newspapers which limits getting FP information (30-32). The poor may also have problems accessing healthcare due to long distances to the health facility, lack of money for transportation, and limited access to FP as a result of out-of-pocket expenditures to purchase FP methods (33).

Our finding found geographical differences to be associated with the FP uptake among women of reproductive age. Specifically, women in Lango sub-region were more likely users of FP methods as compared to other regions of Uganda. This finding is in line with other studies that have also shown geographical differences to influence FP uptake (33-35). This is possible considering some of the interventions by the government and other development agencies operating in the region that has yielded a positive impact with regards to enhancing access to and use of FP methods. For example, the use of voucher plus system ensured that poor women have access to quality maternal health care and FP services at a reduced costs. Furthermore, some regions systematically fail to benefit from wider improvements in health experienced by the general population as such groupings are geographically or linguistically remote, or benefit selectively from national and international investments (36).

The study showed knowledge as a strong predictor of current family planning use. This was also corroborated by Olugbenga et al (2011) in their study carried out in South-Western Nigeria (37). They noted that this pattern should be expected considering much enlightenment that is ongoing on the use of FP in the country. Education exposes women to reproductive health information and empowers them to make appropriate judgments. It is however worth noting that some family planning methods were unpopular among respondents because they were not readily available and relatively more expensive than other methods. These included male sterilization (vasectomy), female sterilization (tubal ligation), lactational amenorrhea, intrauterine device (IUD) levonorgestrel, and vaginal rings.

This study observed that awareness through listening to the local radios was the predominant source of information for FP methods among women. This finding is in tandem with findings elsewhere that have documented the importance of information in influencing FP (20). The use of public media sources like listening to the radio, watching television, and reading newspapers increase the awareness of people on FP methods. This present study did not find any significant association with FP uptake and watching television, reading newspapers, and receiving text messages via mobile phones. This may be so because the majority of women are illiterate and live in rural areas where they have access to local radios but not television, newspapers, and mobile phone. However, to improve FP use, media in all its forms play a major role in influencing the usage of FP methods (38). Information gives women the freedom of choice and can enable them to make better choices of FP methods in addition of having an opportunity to discuss with their spouses.

### Implications of findings

The findings reveals that designing interventions to bring the delivery of FP methods closer to women in the community would increase the uptake as compared to the health facility based administration of methods. Increased uptake of FP methods brings significant health and other benefits. It further offers a range of potential non-health benefits that encompass expanded education opportunities and empowerment for women, and sustainable population growth and economic development of Uganda.

### Strength and limitations of the study

A major strength of this study is that the data used are nationally representative of women of reproductive age in the entire country, Uganda. Nonetheless, some of the limitations of this study include non-sampling errors and sampling errors. Non-sampling errors were a result of mistakes made in implementing data collection and data processing, such as failure to locate and interview the correct household, misunderstanding of the questions on the part of either the interviewer or the respondent, and data entry errors. Although numerous efforts were made during the implementation to minimize this type of error, non-sampling errors were impossible to avoid and difficult to evaluate statistically. Sampling errors, on the other hand, were evaluated statistically. Since it was a population-based study, health facility factors influencing the use of FP methods were not included in the survey.

## Conclusions

Our study shows the prevalence of current FP use in Uganda to be low and a threat to achieving the SDG 3 target of 3.7.1 by 2030. Some of the key factors influencing the use of current FP are; secondary education, highest household wealth index, geographical differences, knowledge, and awareness creation. Therefore, we recommend the promotion of women’s education, awareness through local radios, improving the knowledge of women, and economic empowerment of women to enable them access FP methods. Policymakers need to amend the existing policies to promote the use of FP methods among young women since they are fewer users of FP methods as compared to older women.

## Data Availability

The dataset used is openly available upon permission from the MEASURE DHS website (URL. http://www.dhsprogram.com/data/available.datasets.cmf).

## Supporting information

S1 Fig 1. Map of Uganda showing 15 regions where the 2016 UDHS was implemented (DOC). S1 Fig2. Sample allocation of clusters and households by region and type of residence (DOC).

## Acknowledgments

We appreciate the USAID DHS Programme for granting us the use of the dataset.

## References

1. The Republic of Uganda MoH. Uganda Bureau of Ststistics 2021 statistical abstract. 2021 [cited 2022 July 26th]; Available from: http://library.health.go.ug/publications/statistics/ubos-statistical-abstract-2021.

2. Statistics UBo. Uganda demographic and health survey 2016. UBOS and ICF Kampala, Uganda, and Rockville, Maryland, USA; 2018.

3. WHO. Family Planning/Contraception Keyfacts, 2018 2018; Available from: https://www.who.int/news-room/fact-sheets/detail/family-planning-contraception.

4. Familt Planning/Contraception Methods: Key Facts. WHO [June 2020]; Available from: https://www.who.int/news-room/fact-sheets/detail/family-planning-contraception.

5. Bongaarts J. Trends in fertility and fertility preferences in sub-Saharan Africa: the roles of education and family planning programs. Genus. 2020;76(1):1–15.

6. Asif MF, Pervaiz Z, Afridi JR, Abid G, Lassi ZS. Role of husband’s attitude towards the usage of contraceptives for unmet need of family planning among married women of reproductive age in Pakistan. BMC Women’s Health. 2021;21(1):1–7.

7. Haslegrave M. Sexual and reproductive health and rights in the sustainable development goals and the post-2015 development agenda: less than a year to go. Reproductive health matters. 2014;22(44):102–8.

8. Norheim OF, Jha P, Admasu K, Godal T, Hum RJ, Kruk ME, et al. Avoiding 40% of the premature deaths in each country, 2010–30: review of national mortality trends to help quantify the UN Sustainable Development Goal for health. The Lancet. 2015;385(9964):239–52.

9. Girma Garo M, Garoma Abe S, Dugasa Girsha W, Daka DW. Unmet need for family planning and associated factors among currently married women of reproductive age in Bishoftu town, Eastern Ethiopia. PloS one. 2021;16(12):e0260972.

10. Ntoimo L. Contraception in Africa: Is the global 2030 milestone attainable? African Journal of Reproductive Health. 2021;25(3):9–13.

11. Ackerson K, Zielinski R. Factors influencing use of family planning in women living in crisis affected areas of Sub-Saharan Africa: A review of the literature. Midwifery. 2017;54:35–60.

12. Birhane A, Alemayehu M, Hadush Z, Medhanyie AA, Ahmed M, Mulugeta A. Factors influencing contraceptive use among women of reproductive age from the pastoralist communities of Afar, Ethiopia: a community-based cross-sectional study. The Ethiopian Journal of Health Development. 2018;32(Special Is).

13. Bolarinwa OA, Olagunju OS. Knowledge and factors influencing long-acting reversible contraceptives use among women of reproductive age in Nigeria. Gates Open Research. 2019;3.

14. Davidson AS, Fabiyi C, Demissie S, Getachew H, Gilliam ML. Is LARC for everyone? A qualitative study of sociocultural perceptions of family planning and contraception among refugees in Ethiopia. Maternal and child health journal. 2017;21(9):1699–705.

15. Gebremedhin AY, Kebede Y, Gelagay AA, Habitu YA. Family planning use and its associated factors among women in the extended postpartum period in Addis Ababa, Ethiopia. Contraception and reproductive medicine. 2018;3(1):1–8.

16. Obwoya JG, Wulifan JK, Kalolo A. Factors influencing contraceptives use among women in the Juba city of South Sudan. International Journal of Population Research. 2018;2018.

17. Sserwanja Q, Musaba MW, Mukunya D. Prevalence and factors associated with modern contraceptives utilization among female adolescents in Uganda. BMC women’s health. 2021;21(1):1–7.

18. Demographic U. health survey (UDHS) 2011. Uganda Bureau of Statistics Kampala, Uganda. 2015.

19. Udhs I. Uganda demographic and health survey. Uganda Bureau of Statistics, Kampala Uganda. 2011.

20. Bakibinga P, Matanda DJ, Ayiko R, Rujumba J, Muiruri C, Amendah D, et al. Pregnancy history and current use of contraception among women of reproductive age in Burundi, Kenya, Rwanda, Tanzania and Uganda: analysis of demographic and health survey data. BMJ open. 2016;6(3):e009991.

21. Ouma S, Turyasima M, Acca H, Nabbale F, Obita K, Rama M, et al. Obstacles to family planning use among rural women in Atiak health center IV, Amuru District, northern Uganda. East African medical journal. 2015;92(8):394–400.

22. Organization WH. SDG 3: Ensure healthy lives and promote wellbeing for all at all ages. Sustainable development goals. 2019.

23. Dhs M. Demographic and Health Surveys. Calverton: Measure DHS, ICF International. 2012.

24. Barrow A, Jobe A, Okonofua F. Prevalence and determinants of unmet family planning needs among women of childbearing age in the Gambia: analysis of nationally representative data. Gates Open Research. 2021;4:124.

25. Hamza WS, Darwish MM. Determinants of family planning use among currently married women aged 15-49 years and their partners: a secondary analysis based on the Egypt Demographic and Health Surveys, 2000 and 2008. Egyptian Journal of Community Medicine. 2019;37(3):111–5.

26. Ibrahim HA. Determinants of birth control use among Kenyan women: evidence from DHS-2008-2009. African J Sci Res. 2016;5(1):1–5.

27. Bekele D, Surur F, Nigatu B, Teklu A, Getinet T, Kassa M, et al. Knowledge and attitude towards family planning among women of reproductive age in emerging regions of Ethiopia. Journal of Multidisciplinary Healthcare. 2020;13:1463.

28. Lakew Y, Reda AA, Tamene H, Benedict S, Deribe K. Geographical variation and factors influencing modern contraceptive use among married women in Ethiopia: evidence from a national population based survey. Reproductive health. 2013;10(1):52.

29. Asiimwe JB, Ndugga P, Mushomi J, Ntozi JPM. Factors associated with modern contraceptive use among young and older women in Uganda; a comparative analysis. BMC public health. 2014;14(1):926.

30. Bardaweel SK, Akour AA, Alkhawaldeh A. Impediments to use of oral contraceptives among refugee women in camps, Jordan. Women & health. 2019;59(3):252–65.

31. Bardaweel SK, Akour AA, Kilani M-VZ. Current knowledge, attitude, and patterns of oral contraceptives utilization among women in Jordan. BMC women’s health. 2015;15(1):1–8.

32. Rios-Zertuche D, Blanco LC, Zúñiga-Brenes P, Palmisano EB, Colombara DV, Mokdad AH, et al. Contraceptive knowledge and use among women living in the poorest areas of five Mesoamerican countries. Contraception. 2017;95(6):549–57.

33. Kabagenyi A, Habaasa G, Rutaremwa G. Low contraceptive use among young females in Uganda: does birth history and age at birth have an influence? Analysis of 2011 Demographic and Health Survey. Journal of contraceptive studies. 2016;1(1).

34. Asresie MB, Fekadu GA, Dagnew GW. Contraceptive use among women with no fertility intention in Ethiopia. Plos one. 2020;15(6):e0234474.

35. Kungu W, Agwanda A, Khasakhala A. Trends and determinants of contraceptive method choice among women aged 15-24 years in Kenya. F1000Research. 2020;9(197):197.

36. Walker B. State of the worlds minorities and indigenous peoples 2013. Events of 2012. 2013.

37. Olugbenga-Bello A, Abodunrin O, Adeomi A. Contraceptive practices among women in rural communities in south-western Nigeria. Global Journal of Medical Research. 2011;11(2).

38. Mghweno L, Katamba P. Nyirabavugirije. AM (2017) Influence of Mass Media on Family Planning Methods among Couples in Gashenyi Sector Rwanda. Internal Journal of Multidisciplinary Research and Development.4(6):336–46.

